# Echocardiographic progression of calcific aortic valve disease in patients with preexisting aortic valve sclerosis

**DOI:** 10.1101/2022.12.02.22283035

**Authors:** Jasmin Shamekhi, Carina Uehre, Baravan Al-Kassou, Marcel Weber, Alexander Sedaghat, Atsushi Sugiura, Nihal Wilde, Matti Adam, Verena Veulemans, Malte Kelm, Stephan Baldus, Georg Nickenig, Sebastian Zimmer

**Affiliations:** Heart Center, Department of Medicine II, University Hospital Bonn, Bonn, Germany; Heart Center, Department of Cardiology, University Hospital Cologne, Cologne, Germany; Heart Center, Department of Cardiology, University Hospital Düsseldorf, Düsseldorf, Germany

**Keywords:** Calcific aortic valve disease, CAVD, aortic valve stenosis, aortic calcification

## Abstract

**Objective:** We aimed to evaluate echocardiographic parameters to predict CAVD progression.

**Background:** Calcific aortic valve disease (CAVD) ranges from aortic valve sclerosis (ASc) with no functional impairment of the aortic valve to severe aortic stenosis (AS). It remains uncertain, which patients with ASc are at particular high risk of developing AS.

**Methods:** We included a total of 153 patients with visual signs of ASc and peak flow velocity (Vmax.) below 2.5m/s at baseline echocardiography. Progression of CAVD to AS was defined as an increase of the Vmax. ≥ 2.5m/s with a delta of ≥ 0.1m/s; stable ASc complied with a Vmax. below 2.5m/s and a delta < 0.1m/s. Finally, we compared clinical and echocardiographic parameters between these two groups.

**Results:** The mean age at baseline was 73.5 (± 8.2) years and 66.7% were of male gender. After a mean follow-up of 1463 days, 57 patients developed AS, while 96 patients remained in the ASc group. The AS group showed significantly more calcification (p < 0.001) and thickening (p < 0.001) of the aortic valve cusps at baseline, although hemodynamics showed no evidence of AS in both groups (ASc group: Vmax. 1.6 ± 0.3 m/s versus AS group: Vmax. 1.9 ± 0.3 m/s; p < 0.001). Advanced calcification (OR (95% CI): 4.8 (1.5 − 15.9); p = 0.009) and a cusp thickness > 0.26cm (OR (95% CI): 16.6 (5.4 – 50.7); p < 0.001) were independent predictors for the development of AS.

**Conclusion:** The acquisition of simple echocardiographic parameter may help to identify patients at particular high risk of developing AS.

## Introduction

Calcific aortic valve disease (CAVD) is the most common valvular heart disease in developed countries (1, 2). CAVD ranges from aortic valve sclerosis (ASc) with no functional impairment of the aortic valve to severe aortic valve stenosis (AS). Especially, elderly patients are frequently affected and the prevalence of CAVD is increasing due to global aging and more accurate diagnostic screening methods (3). The initial stage of CAVD is characterized by visual signs of ASc without obstruction of the left ventricular outflow and is present in almost 30% of adults aged over 65 years (4). Severe AS represents the end-stage of CAVD with hemodynamic compromise resulting in shortness of breath, loss of consciousness and chest pain due to an obstruction of the left ventricular outflow. The prevalence of severe AS is about 3% in adults over 75years old (4, 5). So far, there is no therapy available to prevent the progression of CAVD and it remains uncertain, which patients with ASc are at particular high risk of developing AS. In this study, we evaluated the prevalence of CAVD progression in patients with preexistent ASc and assessed echocardiographic parameters to predict disease progression and identify patients at high risk of developing AS.

## Methods

### Study design and patient population

In this study, we compared clinical and echocardiographic parameters of patients with aortic valve sclerosis at baseline, which either developed aortic valve stenosis (mild, moderate or severe) during follow-up echocardiography (AS group), or remained in the preceding stage with stable calcific aortic valve disease (ASc groups). The study design is shown in **Figure 1**.

**Figure 1.**
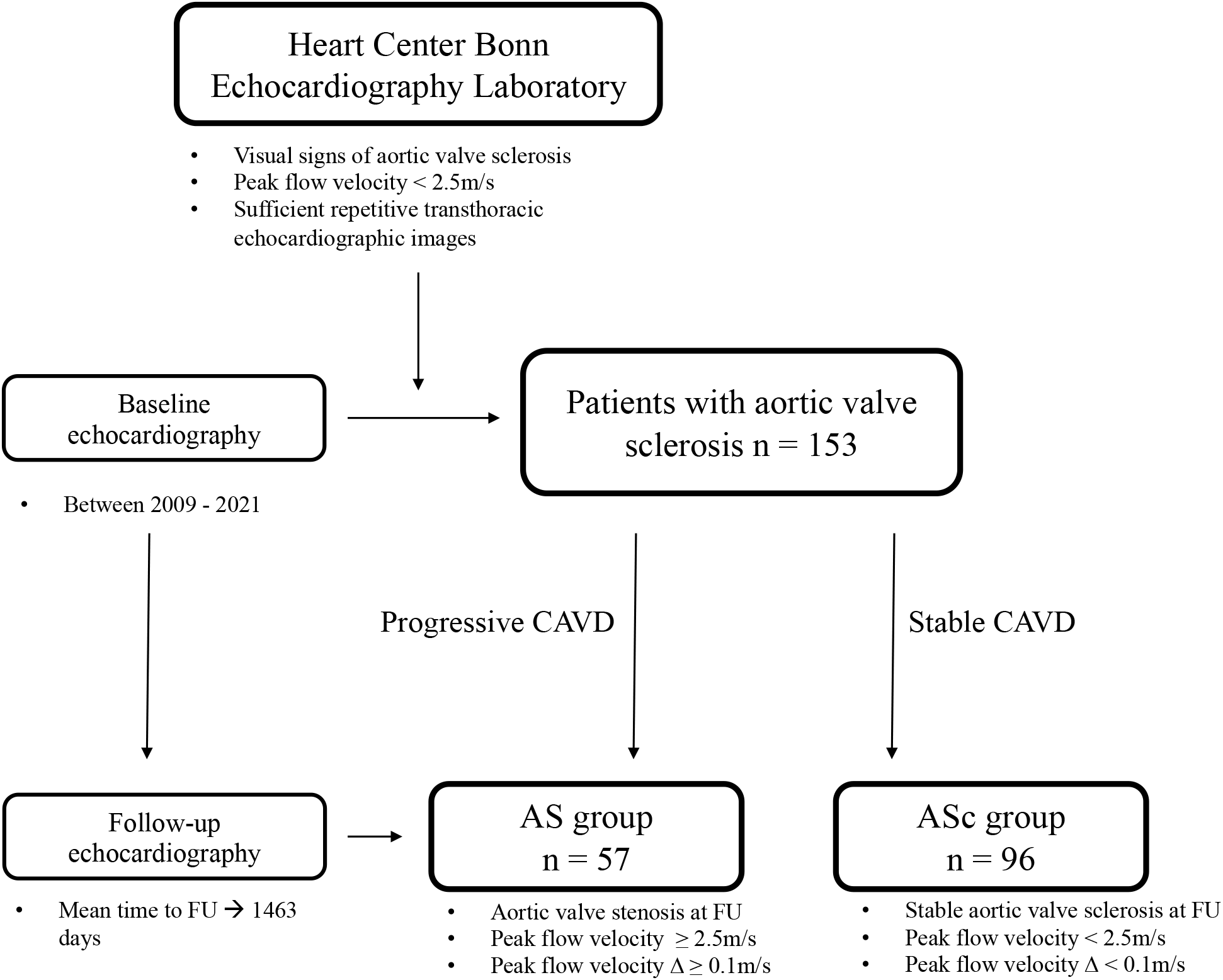
Study design. *AS = aortic stenosis; ASc = aortic sclerosis*

In detail, the database of the echocardiography laboratory of the Heart Center Bonn, which is a consecutive patient data registry, was retrospectively searched for patients with any signs of aortic valve sclerosis without functional impairment of the aortic valve, defined as peak flow velocity below 2.5m/s in transthoracic echocardiography. Prerequisite for the inclusion to the study was the availability of repetitive echocardiographic images (at least two) to evaluate the progression of CAVD over time. Exclusion criteria were missing or incomplete echocardiographic images at baseline or follow-up. Patients with aortic valve prosthesis or bicuspid aortic valve were also excluded from the analysis. The presence of aortic valve sclerosis was assessed by an experienced physician. Progression of CAVD was defined as an increase of the peak flow velocity ≥ 2.5m/s with a delta of at least 0.1m/s (Δ ≥ 0.1m/s); stable CAVD complied with a peak flow velocity below 2.5m/s and a delta < 0.1m/s.

The primary endpoint was the progression of calcific aortic valve disease to any stage of AS. We assessed clinical and echocardiographic parameters including the stage of CAVD at follow-up and stratified patients according to disease progression into two groups: patients with stable calcific aortic valve disease (ASc group) and patients with any stage of aortic valve stenosis (AS group). For the statistical analysis, we compared baseline and echocardiographic parameters between these two groups and evaluated their predictive value to develop aortic valve stenosis.

### Echocardiographic parameters

Transthoracic echocardiography is still the method of choice for the diagnosis and evaluation of aortic valve stenosis (6). The following echocardiographic parameters were assessed and evaluated in this study: left ventricular outflow tract (LVOT) diameter, diameter of the aortic root and the ascending aorta, thickness of the left- (LCC), right- (RCC), and non-coronary cusp (NCC) (measured at the thickest point of the respective cusp), the aortic valve area (AVA) as measured by continuity equation and by planimetry, the mean aortic valve pressure gradient (MPG), the maximum aortic valve pressure gradient (maxPG), the aortic valve peak flow velocity (AV Vmax.), the time to peak velocity, the stroke volume, the systolic duration, the degree of aortic valve regurgitation, visual signs of calcification (divided into minor and major calcification as a binary parameter) and reduced mobility of the left-, right-, and non-coronary cusp (binary variable with the categories “yes” and “no”, respectively), the degree of mitral valve regurgitation, left ventricular hypertrophy, the diastolic und systolic interventricular septal thickness, the degree of diastolic dysfunction, the E/e’ ratio, the left ventricular ejection fraction (LVEF), the left ventricular enddiastolic and endsystolic volume and the left atrial enddiastolic and endsystolic volume. All echocardiographic parameters were assessed in accordance with the recommendations from the American Society of Echocardiography (7).

### Statistical analysis

Data are presented as the mean ± standard deviation, if normally distributed, or as the median and an interquartile range (IQR) (quartile 1/quartile 3), if not normally distributed. Continuous variables were tested for having a normal distribution by using the Kolmogorov-Smirnov test. Categorical variables are given as frequencies and percentages. For continuous variables, a Student’s *t* test or a Mann-Whitney *U* test was performed for comparing between two groups. When comparing more than two groups, ANOVA or the Kruskal–Wallis test was used. Spearman’s correlation coefficients were used to assess associations. The χ^2^ test was used for analysis of categorical variables. To evaluate the prognostic value of the aortic valve cusp thickness for the prediction of disease progression, receiver-operating characteristic (ROC) curves were generated to determine the optimum cut-off value. In consideration of the Youden-Index (Youden-Index = 0.64), a cusp thickness > 0.26cm was used for the statistical analysis. Finally, we performed a multivariate regression analysis, which included univariate predictors with a p-value <0.05, and a ROC curve analysis to assess independent predictors for the progression of CAVD.

Statistical significance was assumed when the null hypothesis could be rejected at p < 0.05. Statistical analyses were conducted with IBM SPSS Statistics version 27.0.0.0 (IBM Corporation, Somers, NY, USA). The investigators initiated the study, had full access to the data, and wrote the manuscript. All authors vouch for the data and its analysis.

## Results

### Overall study population

We identified 153 patients eligible to be included in the study. Clinical and echocardiographic parameters are shown in **Table 1**.

**Table 1.** Clinical and echocardiographic parameters of the overall study population.

| All patients<br>(n = 153) |  |
| --- | --- |
| Clinical parameters |  |
| Age, ± SD (years) | 73.5 ± 8.2 |
| BMI, ± SD (kg/m<sup>2</sup>) | 27.0 ± 4.6 |
| Male sex, n (%) | 102 (66.7) |
| PAD, n (%) | 19 (12.4) |
| CKD, n (%) | 37 (24.2) |
| Dialysis, n (%) | 8 (5.2) |
| Hypertension, n (%) | 132 (86.3) |
| Diabetes, n (%) | 34 (22.2) |
| Dyslipidemia, n (%) | 87 (56.9) |
| Smoker, n (%) | 47 (30.9) |
| Atrial fibrillation, n (%) | 76 (49.7) |
| History of CAD, n (%) | 90 (58.8) |
| Previous stroke, n (%) | 15 (9.8) |
| MAPT, n (%) | 51 (33.6) |
| DAPT, n (%) | 25 (16.3) |
| OAC/DOAC, n (%) | 79 (51.6) |
| Echocardiographic parameters at baseline |  |
| AVA by continuity equation, cm<sup>2</sup> | 1.9 ± 0.7 |
| MPG, mmHg | 6.7 ± 3.0 |
| maxPG, mmHg | 13.0 ± 5.6 |
| Vmax., m/s | 1.7 ± 0.4 |
| Aortic regurgitation, n (%) |  |
| - Grade 0 | 71 (46.4) |
| - Grade I | 65 (42.5) |
| - Grade II | 17 (11.1) |
| - Grade III | - |
| Mitral regurgitation, n (%) |  |
| - Grade 0 | 15 (9.8) |
| - Grade I | 91 (59.5) |
| - Grade II | 42 (27.5) |
| - Grade III | 5 (3.3) |
| Ejection fraction, % | 53.7 ± 11.8 |
| Aortic stenosis, n (%) |  |
| - None | 153 (100) |
| - Mild | - |
| - Moderate | - |
| - Severe | - |
| **Echocardiographic parameters at follow-up** |  |
| Time to follow-up, days | 1463 ± 953 |
| Aortic stenosis at follow-up, n (%) |  |
| - None | 96 (62.7) |
| - Mild | 19 (12.4) |
| - Moderate | 29 (19.0) |
| - Severe | 9 (5.9) |
| AVA by continuity equation, cm<sup>2</sup> | 1.6 ± 0.8 |
| MPG, mmHg | 12.1 ± 9.8 |
| maxPG, mmHg | 23.6 ± 17.7 |
| Vmax., m/s | 2.3 ± 0.8 |
| Aortic regurgitation, n (%) |  |
| - Grade 0 | 67 (43.8) |
| - Grade I | 73 (47.7) |
| - Grade II | 13 (8.5) |
| - Grade III | - |
| Mitral regurgitation, n (%) |  |
| - Grade 0 | 8 (5.2) |
| - Grade I | 93 (60.8) |
| - Grade II | 49 (32.0) |
| - Grade III | 3 (2.0) |
| Ejection fraction, % | 54.5 ± 11.7 |

Values are mean (±SD), median (IQR 1/3) or n/N (%)
AS = aortic stenosis; AScl = aortic sclerosis; BMI = body mass index; PAD = peripheral artery disease; CKD = chronic kidney disease; CAD = coronary artery disease; MAPT = mono antiplatelet therapy; OAC = oral anticoagulant; DOAC = direct oral anticoagulant; AVA = aortic valve area; MPG = mean pressure gradient, maxPG = maximum pressure gradient; AV Vmax. = aortic valve peak flow velocity

The mean age of the overall study cohort was 73.5 ± 8.2 years and 66.7% of the patients were male. Most patients (86.3%) presented with arterial hypertension, 22.2% had diabetes, 56.9% suffered from dyslipidemia and 30.9% were active smokers. Almost two-thirds of the patients had concomitant coronary artery disease. Chronic kidney disease (CKD) was present in 24.2% of the patients, whereof 5.2% had terminal dialysis-dependent renal insufficiency. In the baseline transthoracic echocardiography, the mean AV Vmax. was 1.7 ± 0.4 m/s, the MPG was 6.7 ± 3.0 mmHg and the mean AVA was 1.9 ± 0.7 cm^2^. The mean LVEF was 53.7 ± 11.8 % and 53.6% of the patients suffered from mild to moderate concomitant AR.

The mean time to follow-up was 1463 ± 953 days. At follow-up echocardiography, the mean AV Vmax. of the overall study population was 2.3 ± 0.8 m/s, the MPG was 12.1 ± 9.8 mmHg and the mean AVA was 1.6 ± 0.8 cm^2^. Out of 153 patients, approximately one-third developed AS with a mean AV Vmax. of 3.2 ± 0.5 m/s, whereas 96 patients (63%) showed stable ASc with a mean AV Vmax. of 1.7 ± 0.3 m/s. In detail, 12.4% of the patients developed mild AS, 19.0% showed moderate AS and 5.9% suffered from severe AS.

### Clinical parameters according to CAVD progression

Clinical parameters according to the two CAVD groups (AS vs. ASc) are presented in **Table 2**. The AS group was younger (70.1 ± 10.5 years vs. 75.0 ± 6.0 years; p = 0.001) and presented with higher rates of CKD (35.1% vs. 17.7%; p = 0.01) and dialysis-dependent kidney insufficiency (10.5% vs. 2.1%; p = 0.02) at baseline. Other known risk factors for the development of cardiovascular diseases such as arterial hypertension (p = 0.93), diabetes (p = 0.34), dyslipidemia (p = 0.06) or smoking status (p = 0.11) were not significantly associated with CAVD progression. Both the treatment with oral anticoagulant drugs (p = 0.25) and anti-platelet agents (MAPT: p = 0.52; DAPT: p = 0.09) was not associated with CAVD progression.

**Table 2.** Clinical and echocardiographic parameters according to CAVD progression.

|  | AS group<br>(n = 57) | ASc group<br>(n = 96) | p-value |
| --- | --- | --- | --- |
| **Clinical parameters** |  |  |  |
| Age, ± SD | $70.1 \pm 10.5$ | $75.0 \pm 6.0$ | **0.001** |
| BMI, ± SD | $26.9 \pm 5.4$ | $27.0 \pm 4.1$ | 0.41 |
| Male sex, n (%) | 41 (71.9) | 61 (63.5) | 0.29 |
| PAD, n (%) | 8 (14.0) | 11 (11.5) | 0.64 |
| CKD, n (%) | 20 (35.1) | 17 (17.7) | **0.015** |
| Dialysis, n (%) | 6 (10.5) | 2 (2.1) | **0.023** |
| Hypertension, n (%) | 49 (86.0) | 83 (86.5) | 0.93 |
| Diabetes, n (%) | 15 (26.3) | 19 (19.8) | 0.34 |
| Dyslipidemia, n (%) | 27 (47.4) | 60 (39.2) | 0.06 |
| Smoker, n (%) | 22 (38.6) | 25 (26.3) | 0.11 |
| Atrial fibrillation, n (%) | 26 (45.6) | 50 (52.1) | 0.44 |
| History of CAD, n (%) | 31 (54.4) | 59 (61.5) | 0.39 |
| Previous stroke, n (%) | 8 (14.0) | 7 (4.6) | 0.17 |
| MAPT, n (%) | 17 (30.4) | 34 (35.4) | 0.52 |
| DAPT, n (%) | 13 (22.8) | 12 (12.5) | 0.09 |
| OAC/DOAC, n (%) | 26 (45.6) | 53 (55.2) | 0.25 |
| **Echocardiographic parameters at baseline** |  |  |  |
| LVOT diameter, cm | $2.2 \pm 0.3$ | $2.1 \pm 0.2$ | **0.037** |
| Aortic root diameter, cm | $3.0 \pm 0.4$ | $3.0 \pm 0.3$ | 0.32 |
| Ascending aorta diameter, cm | $2.6 \pm 0.3$ | $2.8 \pm 0.4$ | **<0.001** |
| Cusp thickness NCC, cm | $0.31 \pm 0.06$ | $0.24 \pm 0.05$ | **<0.001** |
| Cusp thickness LCC, cm | 0.29 ± 0.06 | 0.23 ± 0.05 | **<0.001** |
| Cusp thickness RCC, cm | 0.33 ± 0.07 | 0.24 ± 0.05 | **<0.001** |
| AVA plan., cm<sup>2</sup> | 1.7 ± 0.5 | 2.2 ± 0.5 | **<0.001** |
| AVA by continuity equation., cm<sup>2</sup> | 1.7 ± 0.7 | 2.2 ± 0.6 | 0.017 |
| MPG, mmHg | 8.7 ± 3.3 | 5.5 ± 2.0 | **<0.001** |
| maxPG, mmHg | 16.6 ± 5.6 | 10.9 ± 4.4 | **<0.001** |
| Vmax., m/s | 1.9 ± 0.3 | 1.6 ± 0.32 | **<0.001** |
| Time to peak velocity, ms | 94.2 ± 26.0 | 88.8 ± 25.1 | 0.10 |
| Stroke volume, ml | 55.6 ± 18.4 | 57.2 ± 40.7 | 0.39 |
| Systolic duration, sec | 0.3 ± 0.04 | 0.29 ± 0.04 | 0.12 |
| Aortic regurgitation, n (%) |  |  | **<0.001** |
| - Grade 0 | 25 (43.9) | 46 (47.9) |  |
| - Grade I | 21 (36.8) | 44 (45.8) |  |
| - Grade II | 11 (19.3) | 6 (6.3) |  |
| - Grade III | - | - |  |
| Calcification NCC, n (%) | 25 (43.9) | 20 (20.8) | **0.003** |
| Calcification LCC, n (%) | 28 (49.1) | 11 (11.5) | **<0.001** |
| Calcification RCC, n (%) | 44 (78.6) | 27 (28.4) | **<0.001** |
| Calcification anulus, n (%) | 48 (84.2) | 94 (97.9) | **0.002** |
| Calcification leaflet tips, n (%) | 48 (84.2) | 85 (88.5) | 0.44 |
| Reduced mobility NCC, n (%) | 14 (24.6) | 4 (4.2) | **<0.001** |
| Reduced mobility LCC, n (%) | 10 (17.5) | 2 (2.1) | **<0.001** |
| Reduced mobility RCC, n (%) | 16 (28.6) | 8 (8.3) | **<0.001** |
| Mitral regurgitation, n (%) |  |  | 0.60 |
| - Grade 0 | 7 (12.3) | 8 (8.3) |  |
| - Grade I | 30 (52.6) | 61 (63.5) |  |
| - Grade II | 18 (31.6) | 24 (25.0) |  |
| - Grade III | 2 (3.5) | 3 (3.1) |  |
| Heart rate, bpm | 73 (64/87.2) | 68 (60/81.7) | 0.17 |
| LV hypertrophy, n (%) | 31 (54.4) | 54 (56.3) | 0.682 |
| IVSd, cm | 1.3 ± 0.3 | 1.3 ± 0.3 | 0.37 |
| IVSs, cm | 1.6 ± 0.3 | 1.6 ± 0.3 | 0.45 |
| E/e', ± SD | 16.3 (10.8/23.3) | 12.2 (10.2/19.0) | 0.08 |
| Ejection fraction, % | 52.3 (12.2) | 54.5 (11.5) | 0.13 |
| LVEDV, ml | 101.0 (86.8/126.9) | 101.4 (75.2/117.8) | 0.13 |
| LVESV, ml | 47.9 (39.4/58.6) | 47.8 (28.9/58.6) | 0.044 |
| LA volume enddiastolic, ml | 37.9 (22.1/70.1) | 35.6 (22.2/63.8) | 0.69 |
| LA volume endsystolic, ml | 60.6 (41.2/89.5) | 55.5 (41.2/84.9) | 0.54 |
| Diastolic dysfunction, n (%) |  |  | 0.46 |
| - Grade 0 | 20 (35.7) | 33 (34.4) |  |
| - Grade I | 24 (42.9) | 33 (34.4) |  |
| - Grade II | 5 (8.9) | 17 (17.7) |  |
| - Grade III | 7 (12.5) | 13 (13.5) |  |
| **Echocardiographic parameters at follow-up** |  |  |  |
| Time to follow-up, days | 1547 ± 996 | 1414 ± 929 | 0.40 |
| Aortic stenosis, n (%) |  |  | **<0.001** |
| - None | - | 96 (100) |  |
| - Mild | 19 (33.3) | - |  |
| - Moderate | 29 (50.9) | - |  |
| - Severe | 9 (5.9) | - |  |
| LVOT diameter, cm | 2.1 ± 0.3 | 2.1 ± 0.3 | 0.06 |
| Aortic root diameter, cm | 3.0 ± 0.4 | 3.0 ± 0.4 | 0.47 |
| Aortic ascendens diameter, cm | 2.6 ± 0.4 | 2.9 ± 0.4 | **0.001** |
| Cusp thickness NCC, cm | 0.38 ± 0.09 | 0.27 ± 0.06 | **<0.001** |
| Cusp thickness LCC, cm | 0.37 ± 0.08 | 0.25 ± 0.06 | **<0.001** |
| Cusp thickness RCC, cm | 0.40 ± 0.08 | 0.27 ± 0.06 | **<0.001** |
| AVA plan., cm<sup>2</sup> | 0.9 ± 0.4 | 2.0 ± 0.5 | **<0.001** |
| AVA by continuity equation., cm<sup>2</sup> | 1.0 ± 0.3 | 2.1 ± 0.6 | **<0.001** |
| MPG, mmHg | 22.4 ± 8.9 | 5.9 ± 2.4 | **<0.001** |
| maxPG, mmHg | 42.9 ± 14.3 | 13.2 ± 12.4 | **<0.001** |
| Vmax., m/s | 3.2 ± 0.5 | 1.7 ± 0.3 | **<0.001** |
| Time to peak velocity, ms | 98.2 ± 25.9 | 84.4 ± 24.3 | **<0.001** |
| Stroke volume, ml | 53.3 ± 20.0 | 54.7 ± 20.6 | 0.69 |
| Systolic duration, sec | 0.3 ± 0.04 | 0.3 ± 0.05 | 0.16 |
| Aortic regurgitation, n (%) |  |  | 0.14 |
| - Grade 0 | 42 (43.8) | 25 (43.9) |  |
| - Grade I | 49 (51.0) | 24 (42.1) |  |
| - Grade II | 8 (14.0) | 5 (5.2) |  |
| - Grade III | - | - |  |
| Calcification NCC, n (%) | 44 (77.2) | 35 (36.5) | **<0.001** |
| Calcification LCC, n (%) | 49 (86.0) | 26 (27.1) | **<0.001** |
| Calcification RCC, n (%) | 51 (91.1) | 46 (48.4) | **<0.001** |
| Calcification anulus, n (%) | 56 (98.2) | 93 (96.9) | 0.61 |
| Calcification leaflet tips, n (%) | 55 (96.5) | 88 (91.7) | 0.24 |
| Reduced mobility NCC, n (%) | 42 (73.7) | 8 (8.3) | **<0.001** |
| Reduced mobility LCC, n (%) | 35 (61.4) | 4 (4.2) | **<0.001** |
| Reduced mobility RCC, n (%) | 47 (83.9) | 20 (20.8) | **<0.001** |
| Mitral regurgitation, n (%) |  |  | **0.012** |
| - Grade 0 | 6 (10.5) | 2 (2.1) |  |
| - Grade I | 30 (52.6) | 63 (65.6) |  |
| - Grade II | 18 (31.6) | 31 (32.3) |  |
| - Grade III | 3 (5.3) | - |  |
| Heart rate, bpm | 70 (63.5/76) | 68 (58.2/81) | 0.19 |
| LV hypertrophy, n (%) | 44 (77.2) | 57 (59.4) | **0.024** |
| IVSd, cm | 1.4 (0.3) | 1.3 (0.3) | **0.009** |
| IVSs, cm | 1.7 (0.3) | 1.6 (0.3) | 0.08 |
| Ee', ± SD | 16.5 (11.9/23.9) | 15.3 (11/23.6) | 0.20 |
| Ejection fraction, % | 52.6 (13.5) | 55.6 (10.4) | 0.07 |
| LVEDV, ml | 96.6 (76.6/130.2) | 96.7 (72.4/122.9) | 0.59 |
| LVESV, ml | 44.9 (28.0/65.2) | 40.8 (29.5/56.1) | 0.53 |
| LA volume (enddiastolic), ml | 55.4 (32.0/70.9) | 40.3 (25.4/64) | 0.06 |
| LA volume (endsystolic), ml | 70.7 (45.5/97.5) | 62.2 (46.3/82.6) | 0.14 |
| Diastolic dysfunction, n (%) |  |  | 0.12 |
| - Grade 0 | 19 (33.3) | 24 (25.0) |  |
| - Grade I | 25 (43.9) | 32 (33.3) |  |
| - Grade II | 4 (7.0) | 16 (16.7) |  |
| - Grade III | 9 (15.8) | 24 (25.0) |  |

Values are mean ( ± SD), median (IQR 1/3) or n/N (%)
AS = aortic stenosis; ASc = aortic sclerosis; BMI = body mass index; PAD = peripheral artery disease; CKD = chronic kidney disease; CAD = coronary artery disease; MAPT = mono antiplatelet therapy; OAC = oral anticoagulant; DOAC = direct oral anticoagulant; LVOT = left ventricular outflow tract; LCC = left-coronary cusp; RCC = right-coronary cusp; NCC = non-coronary cusp; AVA = aortic valve area; MPG = mean pressure gradient, maxPG = maximum pressure gradient; AV Vmax. = aortic valve peak flow velocity; LV = left ventricular; IVSd = diastolic interventricular septal thickness; IVSs = systolic interventricular septal thickness; LVEDV = left ventricular enddiastolic volume; LVESV = left ventricular endsystolic volume; LA = left atrial

### Echocardiographic parameters according to CAVD progression

Baseline and follow-up echocardiographic parameters according to the CAVD groups are shown in **Table 2**. Patients with CAVD progression (i.e., AS group) presented with a mildly elevated but significantly AV Vmax. (AS group: 1.9 ± 0.3 m/s versus ASc group: 1.6 ± 0.3 m/s; p < 0.001), max PG (AS group: 16.6 ± 5.6 mmHg versus ASc group: 10.9 ± 4.4 mmHg; p < 0.001), and MPG (AS group: 8.7 ± 3.3 mmHg versus ASc group: 5.5 ± 2.0 mmHg; p < 0.001), at baseline. Patients in this group presented with significantly higher rates of major calcification (p < 0.001) and advanced thickening (p < 0.001) of the valve cusps and showed a reduced mobility of the left-coronary (LCC)-, right-coronary (RCC), and non-coronary cusp. Furthermore, the AS group had significantly higher rates of concomitant advanced aortic valve regurgitation (AR) at baseline (AR grade II: 19.3% vs. 6.3; p < 0.001).

At follow-up echocardiography, 19 patients (33.3%) had mild AS, 29 patients (50.9%) presented with moderate AS and 9 patients (5.9%) suffered from severe AS (**Figure 2)**. The mean time to follow-up did not differ between the CAVD groups (AS group: 1547 ± 996 days vs. ASc group: 1414 ± 929 days; p = 0.4). In the AS group, the average MPG was 22.4 ± 8.9 mmHg, the maxPG was 42.9 ± 14.3 mmHg and the mean AV Vmax. was 3.2 ± 0.5 m/s in the follow-up echocardiography. In the ASc group, the average MPG was 5.9 ± 2.4 mmHg, the maxPG was 13.2 ± 12.4 mmHg and the mean AV Vmax. was 1.7 ± 0.3 m/s. A direct comparison between these parameters at baseline and follow-up is demonstrated in **Figure 3**.

**Figure 2.**
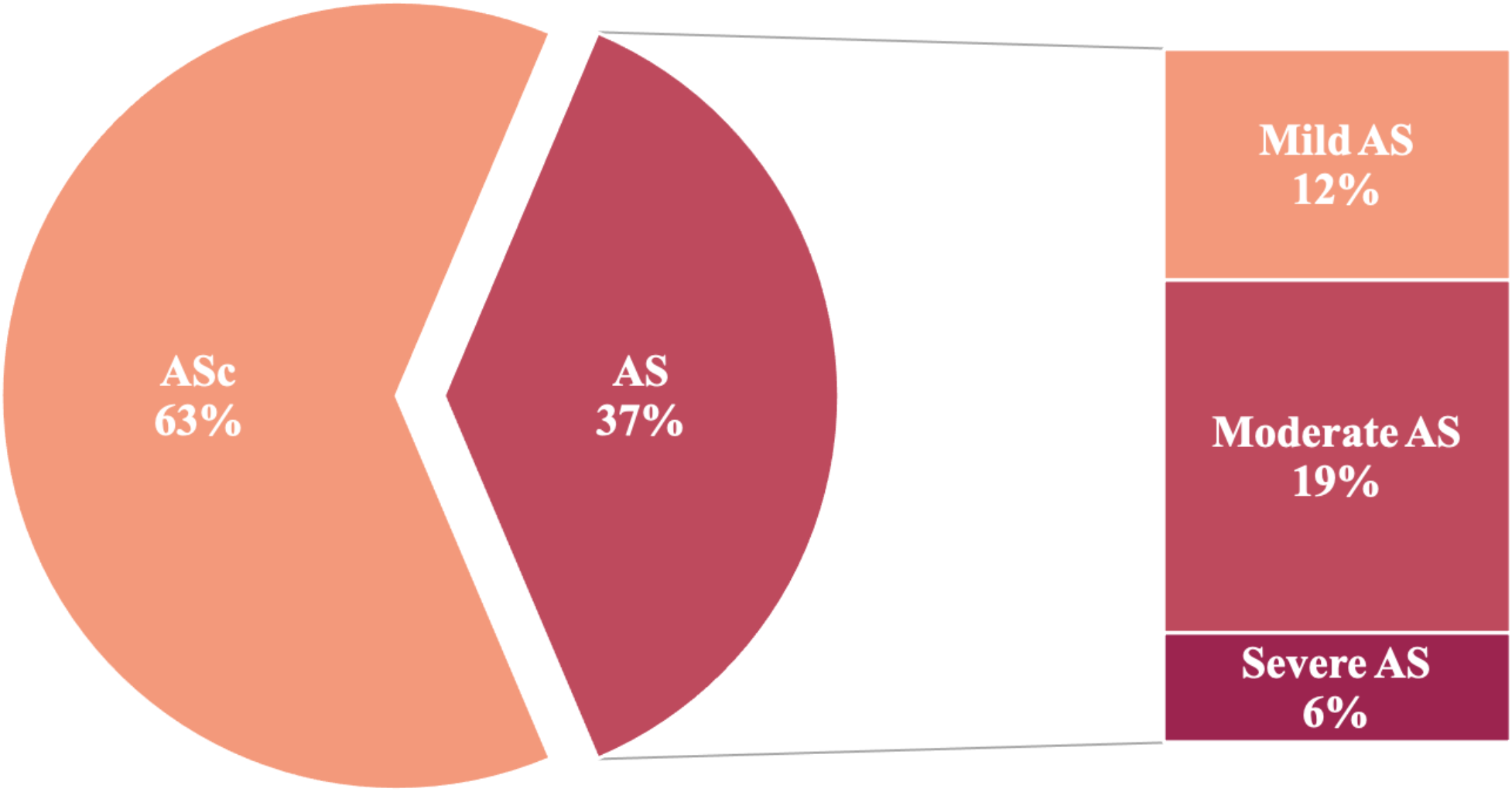
Prevalence of CAVD progression in patients with preexistent aortic valve sclerosis. According to the follow-up echocardiography, 96 (63%) patients showed stable ASc, whereas 57 (37%) of the study patients experienced progression of CAVD; 12.4% of the patients developed mild AS, 19.0% showed moderate AS and 5.9% suffered from severe AS *AS = aortic stenosis; ASc = aortic sclerosis*

**Figure 3.**
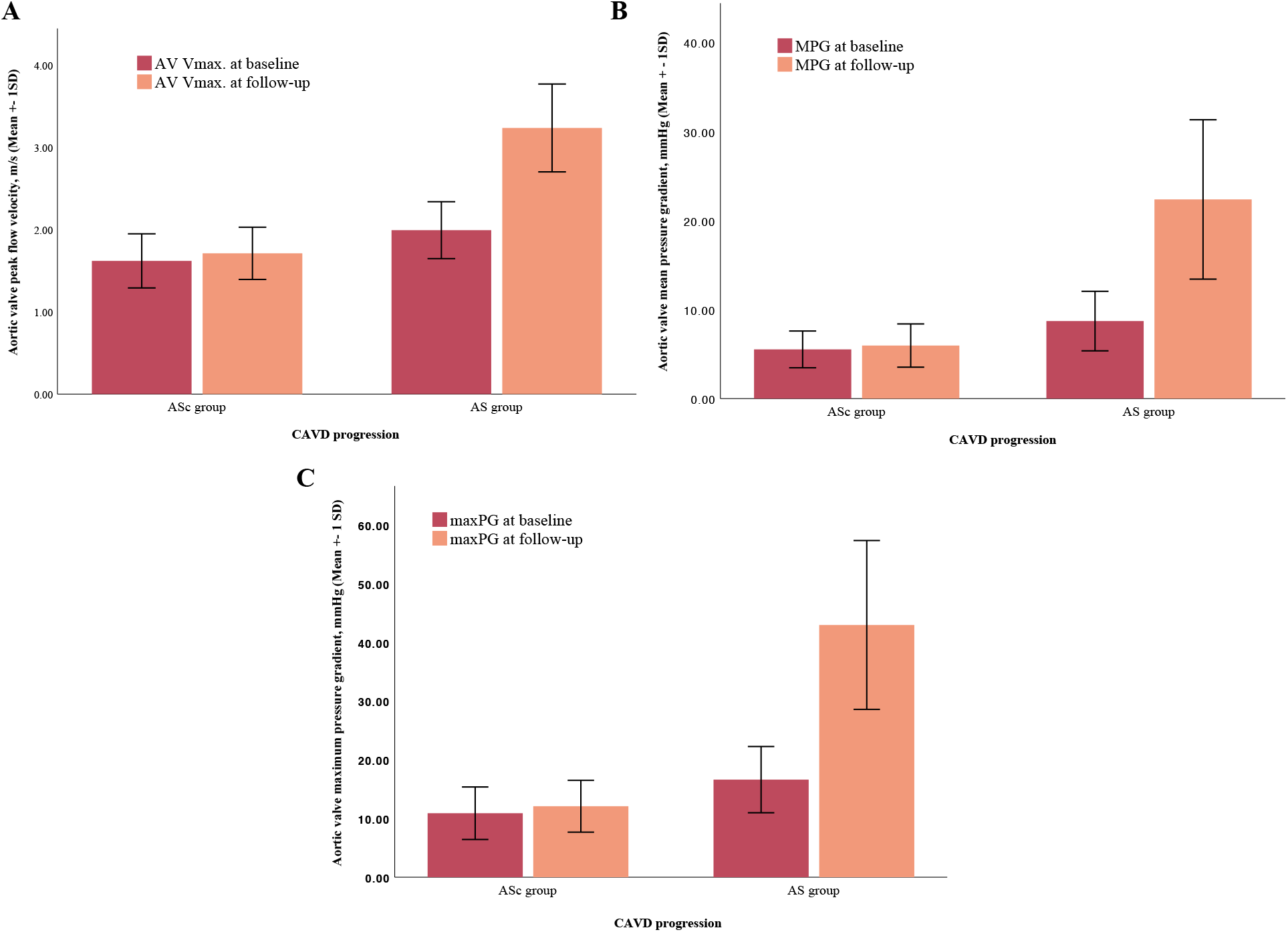
Comparison between echocardiographic parameters at baseline and follow-up in accordance with CAVD progression. The AV Vmax. (**A**), MPG (**B**) and max.PG (**C**) increased significantly within the follow-up period of 4 years in the AS group. *CAVD = calcific aortic valve disease; AS = aortic stenosis; ASc = aortic sclerosis; MPG = mean pressure gradient, maxPG = maximum pressure gradient; AV Vmax. = aortic valve peak flow velocity; SD = standard deviation*

### Multivariate regression analysis

To identify independent predictors for disease progression, we performed a multivariate regression analysis, which included univariate predictors with a p-value <0.05, as shown in **Table 3**. In univariate regression analysis, CKD (p = 0.017), dialysis-dependent kidney insufficiency (p = 0.04), moderate aortic valve regurgitation (p = 0.03), major aortic valve calcification (p < 0.001), reduced valve motion (p < 0.001) and a valve cusp thickness > 0.26cm (p < 0.001) were associated with CAVD progression. The multivariate analysis identified major valve calcification (OR (95% CI): 4.8 (1.5–15.9); p = 0.009) and valve thickness > 0.26cm (OR (95% CI): 16.6 (5.4 – 50.7); p < 0.001) at baseline as independent predictors for the development of AS. CKD (p = 0.06), dialysis-dependent kidney insufficiency (p = 0.19), moderate aortic valve regurgitation (p = 0.5) and reduced valve motion (p = 0.15) were not independently associated with disease progression.

**Table 3.** Multivariate analysis.

| | Univariate analysis<br>HR (95% CI) | $p$ value | Multivariate analysis<br>HR (95% CI) | $p$ value |
| --- | --- | --- | --- | --- |
| Male sex | 2.8 (0.7 – 2.9) | 0.28 | - | - |
| Chronic kidney disease | 2.5 (1.8 – 5.3) | **0.017** | 3.3 (0.9 – 11.8) | 0.06 |
| Dialysis | 5.5 (1.0 – 28.3) | **0.04** | 5.5 (0.4 – 70.6) | 0.19 |
| Ejection fraction | 0.9 (0.9 – 1.1) | 0.25 | - | - |
| Diabetes | 1.4 (0.6 – 3.1) | 0.35 | - | - |
| PAD | 1.2 (0.4 – 3.3) | 0.64 | - | - |
| Atrial fibrillation | 0.8 (0.4 – 1.4) | 0.43 | - | - |
| Arterial hypertension | 0.9 (0.4 – 2.5) | 0.93 | - | - |
| Nicotine abuse | 1.7 (0.8 – 3.5) | 0.11 | - | - |
| Dyslipidemia | 0.5 (0.3 – 1.0) | 0.07 | - | - |
| Moderate aortic regurgitation | 3.3 (1.1 – 10.2) | **0.03** | 1.6 (0.3 – 2.6) | 0.5 |
| Cusp thickness $> 0.26\text{cm}$ | 23.2 (9.2 – 58.5) | **<0.001** | 16.6 (5.4 – 50.7) | **<0.001** |
| Major valve calcification | 13.9 (5.1 – 38.0) | **<0.001** | 4.8 (1.5– 15.9) | **0.009** |
| Reduced valve motion | 6.1 (2.8 – 13.5) | **<0.001** | 2.0 (0.7 – 5.8) | 0.15 |

OR = odds ratio; CI = confidence interval; PAD = peripheral artery disease

### Receiver operating characteristics curve analysis

In a receiver operating characteristics curve analysis, comparing the predictive value of the different echocardiographic parameters for disease progression advanced valve thickness (AUC 0.87 [95% CI: 0.81-0.93], p<0.001) showed the strongest association with disease progression, as presented in **Figure 4**.

**Figure 4.**
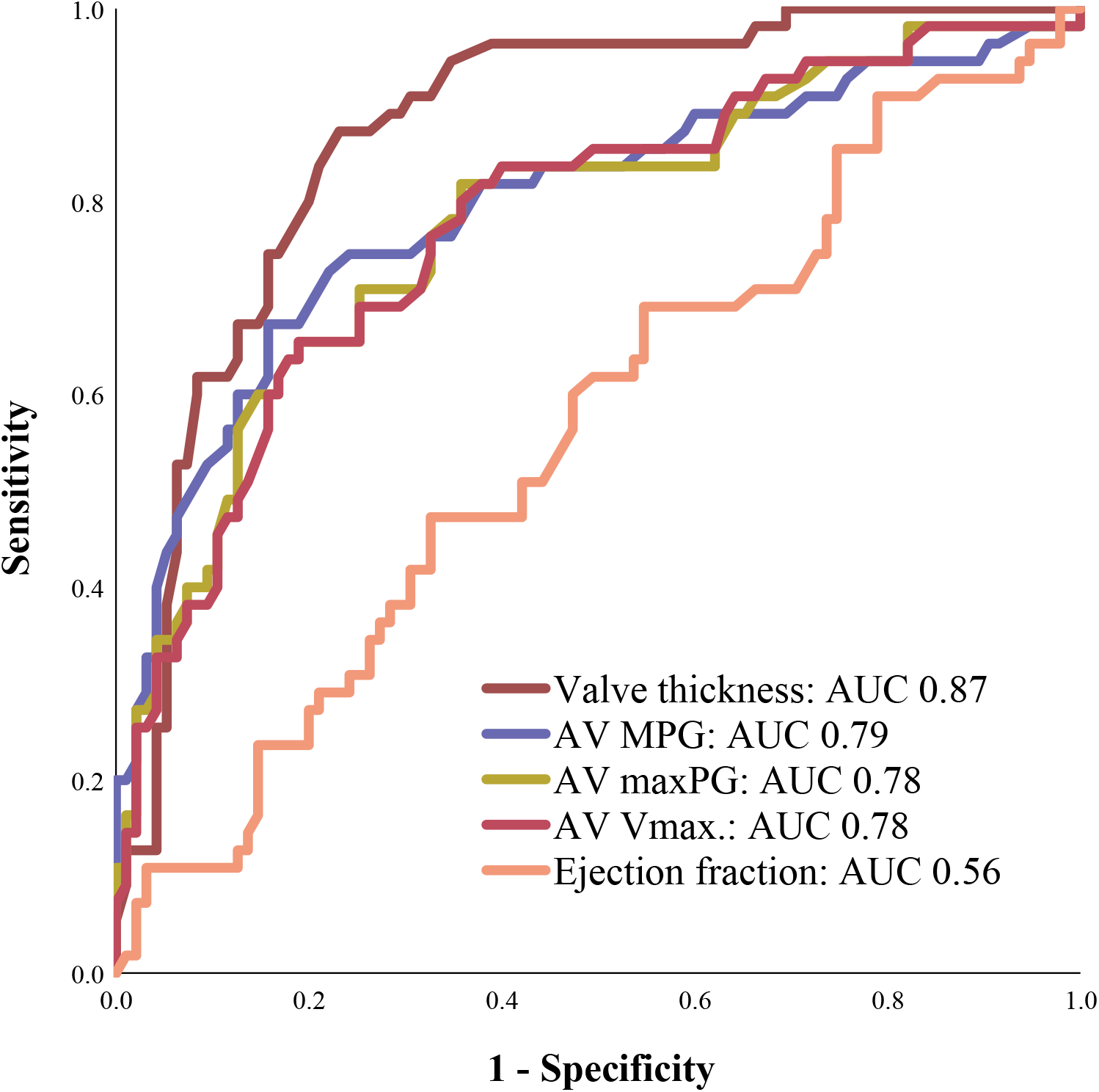
Receiver Operating Characteristics (ROC) curve analysis. Advanced valve thickness showed the strongest association with disease progression in ROC curve analysis. *AUC = area under the curve; MPG = mean pressure gradient, maxPG = maximum pressure gradient; AV Vmax = aortic valve peak flow velocity*

## Discussion

In this study including 153 patients with visual signs of ASc but without AS, we assessed echocardiographic parameters to evaluate the prevalence and the progression of CAVD and to facilitate the identification of patients at high risk to develop aortic valve stenosis. The main results of our study are as follows:

1. Out of 153 study patients, 1/3 experienced progression of CAVD.
2. Traditional cardiovascular risk factors played a minor role in the progression of ASc.
3. Calcification, reduced valve motion and the thickness of the valve cusps were significantly associated with disease progression.
4. Advanced thickness of the valve cusps > 0.26cm and calcification were independent predictors for the development of AS.

### Prevalence of aortic valve sclerosis and disease progression

ASc, the preceding stage of CAVD, is defined as focal areas of valve calcification and leaflet thickening without functional relevant obstruction of the left ventricular outflow tract (8). It is one of the most frequent findings in transthoracic echocardiography with a growing incidence in the older population (9). ASc has been reported to be present in almost 30% of adults aged over 65 years (4, 10), whereas the prevalence of disease progression from ASc to AS differs in the literature. One of the largest prospective studies included > 2000 patients with ASc and a mean age of 69 years, of whom 16% developed AS within 8 years of follow-up; 10.5% developed mild stenosis, 3% had moderate stenosis and 2.5% suffered from severe aortic stenosis (11). Contrary, a meta-analysis of twenty-two studies revealed a progression rate of 1.8-1.9% of patients per year in individuals with baseline ASc (8). *Faggiano et al*. found a progression rate from ASc to some degree of AS of 32.7% in a smaller cohort of 400 patients with a mean age of 68 years during a follow-up period of 4 years; 2.5% of the patients developed severe AS, 5.2% processed to moderate AS and 25% experienced mild AS. Comparable results could be observed in our study. We found a progression rate of 37% within 4 years of follow-up in a cohort of 153 patients with a mean age of 73 years; 12% of patients developed mild AS, 19% presented with moderate AS and 6% progressed to severe AS. But despite the high prevalence of CAVD and its clinical implication, we are still not able to break the circle and to delay or even prevent disease progression. An unfortunate fact, considering the lack of any form of pharmacological treatment in patients with hemodynamically relevant AS.

### CAVD and comorbidities

Several studies have already evaluated the overlap of traditional cardiovascular risk factors (CRF) and the presence of aortic valve calcification (12-16). In the past, comorbidities such as advanced age, male gender, arterial hypertension, dyslipidemia and smoking have been shown to be associated with the development of aortic valve calcification and atherosclerotic disease to a comparable degree (12, 17), supporting the hypothesis that both diseases have a shared developing process. These data are supported by our study results, as we observed a high prevalence of CRF and concomitant coronary artery disease at baseline in our study population. However, with regard to CAVD progression, the presence of traditional CRF seemed to play only minor role, as we could not determine a significant association between progressive disease and rates of CRF. Similar findings have been described by *Messika-Zeitoun et al*. on the basis of a prospective analysis including 70 patients with baseline aortic valve calcification. The study results showed that the progression of established ASc was unrelated to cardiovascular risk factors, age and gender (18). *Bellamy et al*. evaluated the association between CAVD progression and cholesterol levels at baseline in a cohort of 156 patients without revealing a significant correlation between blood cholesterol concentrations and the progression of ASc (19). Corroborating results have been described by other major prospective studies including SEAS, SALTIRE and ASTRONOMER. These trials could not find a relationship between LDL levels and progressive aortic valve disease on the one hand, and were not able to confirm the beneficial effect of statins on CAVD progression, on the other (20-22). In our study, chronic kidney disease (CKD) and terminal dialysis-dependent renal insufficiency were the only clinical factors, that were significantly associated with disease progression. This result is not surprising, as CKD and especially long-term dialysis are often linked with the occurrence of cardiovascular events. Interestingly, patients with CAVD progression were significantly younger with a mean age of 70 years at baseline compared to patients with stable ASc, who were, on average, five years older. Summarizing, our results suggest that other factors than the known CRF have to be involved in the progression of aortic valve calcification. Larger prospective studies are needed to identify these factors, to ensure the development of targeted therapies.

### Transthoracic echocardiography and CAVD

Another important goal should be the reliable and early identification of patients with ASc, who are at high risk to develop AS. In this context, imaging techniques play an important role. Transthoracic echocardiography (TTE) is the gold standard for the evaluation of CAVD and the quantification of AS severity. Beside the visual assessment of the leaflet anatomy and the extend of valve calcification, the evaluation of functional parameters are pivotal in the diagnostic work-up (23, 24).

In our study, we evaluated echocardiographic parameters with regard to their forecast value to predict the development of AS and identified the degree of calcification, valve thickening and reduced valve motion to be associated with CAVD progression. In multivariate analysis, major calcification and valve thickness > 0.26cm were independent predictors for the development of AS. However, one must be aware that the reliable and accurate identification of aortic valve calcification using echocardiography is still challenging given the variability of scanner settings, image quality and the examiners’ experience. In our study, the quantification of ASc based on visual assessment, as a precise and objective quantification of aortic valve sclerosis in the early-stage is due to the limited resolution of the TTE nearly impossible. The only alternative to quantify aortic valve sclerosis more precisely would be the examination by computed tomography (25). Which unfortunately would be accompanied by a high radiation exposure, especially if repetitive examinations are needed. Hence, the huge advantage of the assessment of the visual echocardiographic parameters described above, is the simple, non-invasive, cost-effective and radiation-free acquisition, which could be performed easily in every routine TTE examination. As a consequence, patients with ASc and visual signs of advanced calcification and valve thickening, could be closely monitored with regard to echocardiographic signs of disease progression and the new onset of symptoms.

## Study limitations

Major limitations of our study are the sample size and the retrospective character, representing an important selection bias; the study patients were referred to the transthoracic echocardiography for various indications and we cannot exclude that the reason for the follow-up examination was a symptomatic AS in some patients. Furthermore, we have no survival or functional outcome data available, so that the impact of CAVD progression on outcome in our study population remains uncertain. Therefore, the results of this study should be considered hypothesis-generating. Prospective and larger trials are necessary to confirm our results.

## Conclusion

One-third of patients with aortic valve sclerosis at baseline developed some degree of AS within a follow-up period of four years. Advanced aortic valve calcification and a cusp thickness > 0.26cm at baseline echocardiography were independent predictors for the development of AS in these patients (**Central Illustration**). The acquisition of simple echocardiographic parameter can help to identify patients at particular high risk to develop aortic valve stenosis.

**Central Illustration.**
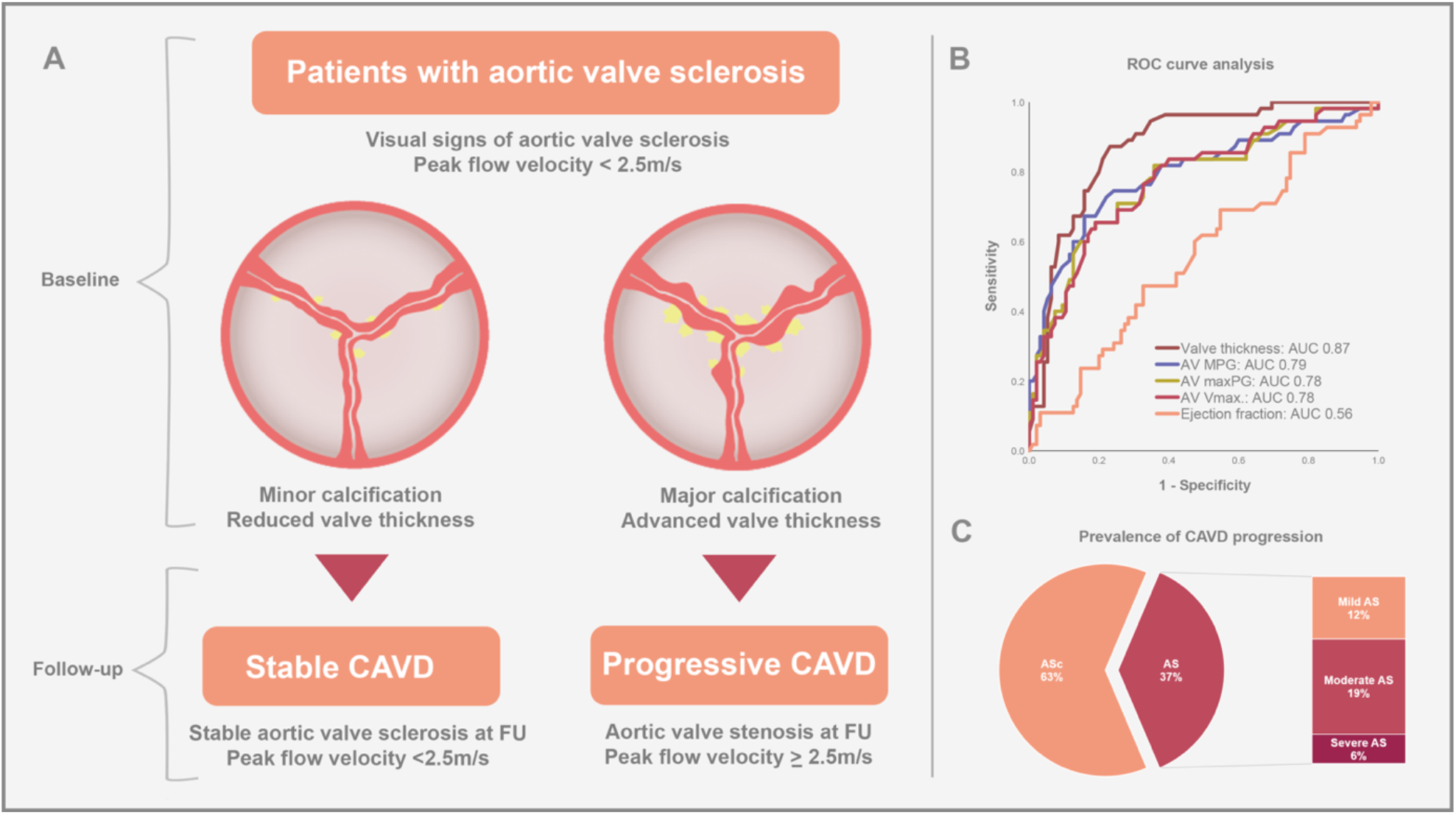
Progression of calcific aortic valve disease in patients with preexisting aortic valve sclerosis within a follow-up period of 4 years. **A** Major aortic valve calcification and thicker valve cusps at baseline echocardiography are independent predictors for the development of AS. **B** Receiver Operating Characteristics (ROC) curve analysis. Advanced valve thickness showed the strongest association with disease progression in ROC curve analysis. **C** Prevalence of CAVD progression in patients with preexistent aortic valve sclerosis. At follow-up echocardiography, 96 (63%) patients showed stable ASc, whereas 57 (37%) of the study patients experienced progression of CAVD; 12.4% of the patients developed mild AS, 19.0% showed moderate AS and 5.9% suffered from severe AS *AS = aortic stenosis; ASc = aortic sclerosis; AUC = area under the curve; CAVD = calcific aortic valve disease; FU = follow-up; MPG = mean pressure gradient, maxPG = maximum pressure gradient; AV Vmax. = aortic valve peak flow velocity*

## Data Availability

All data produced in the present work are contained in the manuscript

## Abbreviation List

CAVD: Calcific aortic valve disease
ASc: Aortic valve sclerosis
AS: Aortic valve stenosis
AVA: aortic valve area
MPG: mean pressure gradient
maxPG: maximum pressure gradient
AV Vmax.: aortic valve peak flow velocity
LCC: left-coronary cusp
RCC: right-coronary cusp
NCC: non-coronary cusp
LVOT: left ventricular outflow tract
AR: aortic valve regurgitation
CKD: chronic kidney disease
COPD: Chronic obstructive pulmonary disease
LVEF: Left ventricular ejection fraction

## References

1. Iung B, Vahanian A. Degenerative calcific aortic stenosis: a natural history. Heart 2012; 98:7–13.

2. Zheng KH, Tzolos E, Dweck MR. Pathophysiology of Aortic Stenosis and Future Perspectives for Medical Therapy. Cardiol Clin. 2020; 38:1–12.

3. Peeters FECM, Meex SJR, Dweck MR, Aikawa E, Crijns HJGM, Schurgers LJ, Kietselaer BLJH. Calcific aortic valve stenosis: hard disease in the heart: A biomolecular approach towards diagnosis and treatment. Eur Heart J. 2018; 39:2618–2624.

4. Beckmann E, Grau JB, Sainger R, Poggio P, Ferrari G. Insights into the use of biomarkers in calcific aortic valve disease. J Heart Valve Dis. 2010; 19:441–52.

5. Small A, Kiss D, Giri J, Anwaruddin S, Siddiqi H, Guerraty M, Chirinos JA, Ferrari G, Rader DJ. Biomarkers of Calcific Aortic Valve Disease. Arterioscler Thromb Vasc Biol. 2017; 37:623–632.

6. Alec Vahanian, Friedhelm Beyersdorf, Fabien Praz, Milan Milojevic, Stephan Baldus, Johann Bauersachs, Davide Capodanno, Lenard Conradi, Michele De Bonis, Ruggero De Paulis, Victoria Delgado, Nick Freemantle, Kristina H Haugaa, Anders Jeppsson, Peter Jüni, Luc Pierard, Bernard D Prendergast, J Rafael Sádaba, Christophe Tribouilloy, Wojtek Wojakowski. 2021 ESC/EACTS Guidelines for the management of valvular heart disease. EuroIntervention. 2021 Dec 22.

7. Mitchell C, Rahko PS, Blauwet LA, Canaday B, Finstuen JA, Foster MC, Horton K, Ogunyankin KO, Palma RA, Velazquez EJ. Guidelines for Performing a Comprehensive Transthoracic Echocardiographic Examination in Adults: Recommendations from the American Society of Echocardiography. J Am Soc Echocardiogr. 2019; 32:1–64.

8. Coffey S, Cox B, Williams MJ. The prevalence, incidence, progression, and risks of aortic valve sclerosis: a systematic review and meta-analysis. Am Coll Cardiol. 2014; 63:2852–61.

9. Faggiano P, Antonini-Canterin F, Erlicher A, et al. Progression of aortic valve sclerosis to aortic stenosis. Am J Cardiol. 2003; 91:99–101.

10. Freemann RV, Otto CM. Spectrum of calcific aortic valve disease: pathogenesis, disease progression, and treatment strategies. Circulation. 2005; 111:3316–26.

11. Cosmi JE, Kort S, Tunick PA, Rosenzweig BP, Freedberg RS, Katz ES, Applebaum RM, Kronzon I. The risk of the development of aortic stenosis in patients with “benign” aortic valve thickening. Arch Intern Med. 2002; 162:2345–7.

12. Stewart BF, Siscovick D, Lind BK, Gardin JM, Gottdiener JS, Smith VE, Kitzman DW, Otto CM. Clinical factors associated with calcific aortic valve disease: Cardiovascular Health Study. J Am Coll Cardiol. 1997; 29: 630–634.

13. Otto CM, Lind BK, Kitzman DW, Gersh BJ, Siscovick DS. Association of aortic-valve sclerosis with cardiovascular mortality and morbidity in the elderly. N Engl J Med. 1999; 341: 142–147.

14. Aronow WS, Schwartz KS, Koenigsberg M. Correlation of serum lipids, calcium, and phosphorus, diabetes mellitus and history of systemic hypertension with presence or absence of calcified or thickened aortic cusps or root in elderly patients. Am J Cardiol. 1987; 59: 998–999.

15. Lindroos M, Kupari M, Valvanne J, Strandberg T, Heikkila J, Tilvis R. Factors associated with calcific aortic valve degeneration in the elderly. Eur Heart J. 1994; 15: 865–870.

16. Boon A, Cheriex E, Lodder J, Kessels F. Cardiac valve calcification: characteristics of patients with calcification of the mitral annulus or aortic valve. Heart. 1997; 78: 472–472.

17. Wilson PWF, D’Agostino RB, Levy D, Belanger AM, Silbershatz H, Kannel WB. Prediction of coronary heart disease using risk factor categories. Circulation. 1998; 97: 1837–1847.

18. Messika-Zeitoun D, Bielak LF, Peyser PA, et al. Aortic valve calcification: determinants and progression in the population. Arterioscler Thromb Vasc Biol. 2007; 27:642–8.

19. Bellamy MF, Pellikka PA, Klarich KW, Tajik AJ, Enriquez-Sarano M. Association of cholesterol levels, hydroxymethylglutaryl coenzyme-A reductase inhibitor treatment, and progression of aortic stenosis in the community. J Am Coll Cardiol 2002; 40:1723–30.

20. Cowell SJ, Newby DE, Prescott RJ, et al. A randomized trial of intensive lipid-lowering therapy in calcific aortic stenosis. N Engl J Med 2005; 352:2389–97.

21. Rosseo AB, Pedersen TR, Boman K, et al. Intensive lipid lowering with simvastatin and ezetimibe in aortic stenosis. N Engl J Med 2008; 359:1343–56.

22. Chan KL, Teo K, Dumesnil JG, et al. Effect of Lipid lowering with rosuvastatin on progression of aortic stenosis: results of the aortic stenosis progression observation: measuring effects of rosuvastatin (ASTRONOMER) trial. Circulation 2010; 121:306–14.

23. Vahanian A, Beyersdorf F, Praz F, et al. 2021 ESC/EACTS Guidelines for the management of valvular heart disease. Eur Heart J. 2021 Aug 28: ehab395.

24. Otto C, Pearlman AS, Gardner CL. Hemodynamic progression of aortic stenosis in adults assessed by Doppler Echocardiography. J Am Coll Cardiol. 1989: 13545–13550.

25. Rajamannan NM, Evans FJ, Aikawa E, et al. Calcific Aortic Valve Disease: Not Simply a Degenerative Process A Review and Agenda for Research from the National Heart and Lung and Blood Institute Aortic Stenosis Working Group. Circulation. 2011; 124:1783–1791.

